# CHANGE IN STROKE SURVIVAL in Sweden 2000 – 2022 – the importance of sex, attained education and age

**DOI:** 10.64898/2026.08.27.26361579

**Authors:** Kristian Bolin, Katharina S Sunnerhagen

**Author notes:** Correspondence: Kristian Bolin, Department of Economics, Centre for Health Governance, University of Gothenburg, PO Box 640, S-405 30 Gothenburg, Sweden.

## Abstract

**Background:** The time trend in long-term survival after a stroke is to some extent unknow due to (relatively) short follow up periods in available data. The objective of this study is to identify and quantify differences in long-term stroke survival in Sweden between men and women and patients with different attained educational levels, comparing two time-periods, 2000-2009 and 2010-2022.

**Methods:** This study employs total population Swedish register data pertaining to hospital-based care and mortality due to stroke for the period 2000–2022 in order to estimate survival (all-cause mortality) after ischaemic and haemorrhagic stroke, respectively, and pertaining to attained educational level. Kaplan-Meier survival functions are estimated stratifying for time-period, sex and educational level. Cox regressions are employed to quantify mortality hazard ratios between the strata. Age is taken into account in complementary analyses (supplement).

**Results:** Taking only time-period (2000-2009 vs 2010-2022) into account resulted in significantly higher survival in the second period for ischaemic stroke patients (HR: 0.84; 95% CI: 0.83-0.84), while no significant difference could be detected for haemorrhagic stroke. Stratifying for sex showed that men gained more than women in terms of reduced mortality hazard rate between the periods. Further stratifying by educational level and estimating survival separately for men and women showed that, for both men and women, patients with the lowest education were relatively worse off (compared to patients with higher education) in the second period. Further analyses, taking age into account, reversed the relative hazard ratio between men and women, but corroborated the result that low education is associated with poorer outcome than high education.

**Conclusions:** The results suggest that there are considerable differences in expected long-term survival after stroke between the sexes, but that this may be due to differences in age between the sexes at the time of stroke. Moreover, lower educational level is significantly associated with lower long-time survival.

## INTRODUCTION

While stroke mortality rates have declined over the last decades in most countries, it is still a major cause of mortality and morbidity entailing substantial economic costs and diminishing quality of life (Feigin et al., 2024; Stark et al., 2021; Yu et al., 2025; Xie et at., 2025). Long-term survival (> 3 years) of stroke patients is less studied (Peng et al., 2022; Aked et al., 2021), and, in particular, *the trend* in long term survival is largely unknown. This paper is based on this observation and findings or conjectures emerging in the recent literature: (1) the decline in stroke mortality is confined to ischaemic stroke (Drescher at al., 2026; Wolsink et al., 2025); (2) the decline in mortality, or lack thereof in case of hemorrhagic stroke, reflects the *joint* effect of enhances in treatment (the introduction of reperfusion therapy and a greater range of anticoagulant drugs), improved diagnostics, and enhanced prevention – e.g., oral anticoagulants (Drescher et al., 2026; Berge et al., 2021; 2021; Sarraj et al., 2023); (3) there are significant differences in stroke mortality and mortality trends between socioeconomic groups and between the sexes, and there is emerging evidence suggesting that the advances in stroke treatment have not benefited or been made available to all groups in society equally (or at all, in some cases) (Apostolaki-Hansson et al., 2025; Bray et al., 2018; Yu et al., 2025; Pantoja-Ruiz et al., 2024; Lindmark et al., 2020); (4) Despite the aforementioned improvements in treatments available to stroke patients during the last decades our understanding of the long-term health effects of stroke comes from studies with short follow-up time (Peng et al., 2022).

The extent to which each specific of the aforementioned factors has contributed to the decline in mortality among (ischaemic) stroke patients is unclear (Drescher et al., 2026). Moreover, published evidence is not decisive as to whether mortality among (ischaemic) stroke patients has continued to decrease after (about) 2010 (Feigin et al., 2009; Wafa et al., 2020; Yafasova et al., 2020; Zhang et al., 2020; Renedo et al., 2024), although it is recognized that the length of time periods covered may affect conclusions about (stroke) mortality trends (Drescher et al., 2026).

Thus, based on the above, this study analyzes the trend in long-term survival following a stroke, distinguishing between different types of stroke and different time periods, sex and educational attainment (stroke before versus stroke after a specific point in time, employing Swedish total-population individual-level register data for the years 2000 - 2022). The cut-off point in time is set at 2009-12-31. This is not perfect in the sense that neither of the new treatment and prevention options were available prior to 2010, but the significant increase in the use of reperfusion therapy came around 2008 – 2010 (in Sweden), and non-vitamin K oral anticoagulants (NOACs) were introduced in Sweden after 2010.

## DATA AND METHODS

The data employed in this study was collected from Swedish total-population individual-level registers and comprise information about hospitalizations and mortality for the years 2000 – 2022. More specifically, the study population was identified by the following criteria: at least one hospitalization registered in the Swedish Patient Registry (Swedish National Board of Health and Welfare) with main diagnosis ICD10: I61 (intracerebral haemorrhage), I63 (cerebral infarction) or I64 (unspecified stroke). The information collected includes date of hospital admission, date of discharge, registered main diagnosis, date of death and (main) cause of death according to ICD10 (Swedish National Board of Health and Welfare). This data was complemented with information about attained education (Statistics Sweden; primary school, high school, post-high school and post graduate school) and collapsed into two categories: high school and post high school.

### Data analysis

The study period 2000 – 2022, was dived in the two aforementioned periods: 2000 – 2008 and 2009 – 2022 respectively, thus distinguishing between (a first observed) stroke in the first and second period. These two populations are described by absolute frequencies of hospitalized patients and (average) absolute incidence rates (calculated using demographic statistics, Statistics Sweden) by type of stroke (ischaemic and haemorrhagic), time-period and sex. The populations are further stratified in age groups (<40, 40 – 50, 50 – 60, 60 – 70, 70 – 80, 80 – 90, > 90) for which the distribution of each population is reported. Educational level (< highs school, >= highs school) is reported as share of patients with high school education or beyond. Two types of survival analyses were performed. Kaplan-Meier survival functions were estimated comparing the two time periods, and sex and educational level (Log-Rank test were computed to infer the statistical differences between Kaplan-Meier survival curves). In order to quantify differences in survival between compared categories, parametric hazards functions were estimated using Cox regressions.

Survival durations were calculated as time (days) between first observed hospitalization due to ischaemic and haemorrhagic stroke, respectively, and date of death or (censoring) date of study end (31^st^ of December 2022). Four Kaplan-Meier survival functions were estimated for each of the two types of strokes, comparing long-term survival between time periods, between the sexes and between educational groups. Correspondingly, hazard functions were estimated by Cox regressions quantifying the hazard ratios between aforementioned groups. Cox regression results (hazard ratios comparing (all-cause) mortality rates between time periods) involving a further stratification by age are reported without illustrating the corresponding Kaplan-Meier survival functions (these are reported in the online supplement).

## Result

Frequencies of hospitalized patients by type of stroke are reported in Table 1, separately for the two time periods, 2000 – 2009 and 2010 – 2022, i.e., a 9-year and a 12-year follow-up period, respectively. Age- and educational distributions are reported as shares of male and female populations in each age group (<50, 50 - < 80, >= 80), and shares having an educational attainment beyond high school. A total of 95 164 (96 340) male (female) patients were hospitalized due to ischaemic stroke in the period 2000 – 2008 (first time-period). The corresponding number in the period 2009 – 2022 (second time-period) were 108 610 (99 868). Average absolute incidence rates of ischaemic stroke declined from the first to the second time-period, for both men and women, whereas the corresponding figures for homorrhagic stroke were roughly the same in both periods. The age distributions show some changes over the two time-periods. In particular, the shares of both the male and female ischaemic stroke population who had their first stroke before 50 years of age increased. The same pattern can be seen for hemorrhagic stroke.

**Table 1.** Aggregate statistics for the period 2000 – 2022, by time-period, type of stroke and sex. Number of unique patients hospitalized due to ischaemic stroke, and hemorrhagic stroke, respectively, corresponding average absolute incidence rates, age distribution, and educational attainment.

|  |  |  |  |  |  |
| --- | --- | --- | --- | --- | --- |
|  | Time period: <= 2009 |  |  | Time period: >2009 |  |
| Ischaemic stroke |  |  |  |  |  |
|  | Men | Women |  | Men | Women |
| # Patients / absolute incidence rate |  |  |  |  |  |
|  | 95 164 (0.21%) | 96 340 (0.21%) |  | 108 610 (0.17%) | 99 868 (0.15%) |
| Share >= high school |  |  |  |  |  |
|  | 51% | 43% |  | 63% | 57% |
| Age distribution |  |  |  |  |  |
| <50 | 3% | 2% |  | 4% | 3% |
| 50 - < 80 | 62% | 44% |  | 63% | 45% |
| >= 80 | 35% | 54% |  | 33% | 52% |
| Hemorrhagic stroke |  |  |  |  |  |
| # Patients / absolute incidence rate |  |  |  |  |  |
|  | 17 011 (0.04%) | 14 676 (0.03%) |  | 20 659 (0.03%) | 17 390 (0.03%) |
| Share >= high school |  |  |  |  |  |
|  | 56% | 49% |  | 65% | 61% |
| Age distribution |  |  |  |  |  |
| <50 | 8% | 5% |  | 8% | 6% |
| 50 - < 80 | 66% | 49% |  | 62% | 47% |
| >= 80 | 26% | 45% |  | 30% | 47% |

### Change in survival over time – ischaemic stroke

The four estimated Kaplan-Meier survival functions are illustrated in Figures 1 – 4. Figure 1 compares survival (days) between first-period and second-period ischaemic stroke patients. Survival among patients having their first ischaemic stroke after 2009 is significantly higher than the corresponding survival among patients before 2010 (log-rank test *p* = 0.000). A Cox regression estimated the corresponding hazard ratio (HR) at 0.84 (95% CI: 0.83-0.84) (second time-period vs first time-period). Stratifying by sex leads to four estimated survival functions illustrated in Figure 2 (log-rank test men *p* = 0.000, log-rank women *p* = 0.000). Figure 2 demonstrates that the difference in survival between the time periods is qualitatively the same for both sexes. However, there are quantitative differences: the HR for men was estimated at 0.82 (95% CI: 0.81-0.83) and the correspond HR for women at 0.87 (95% CI: 0.86-0.88). Cox regressions separately for men and women (time period as explanatory variable) show that the difference between the HR’s of men and women comparing the two time periods is statistically significant inferred from non-overlapping (95 % CIs: 0.85 – 0.88; 0.81 – 0.83).

**Figure 1.**
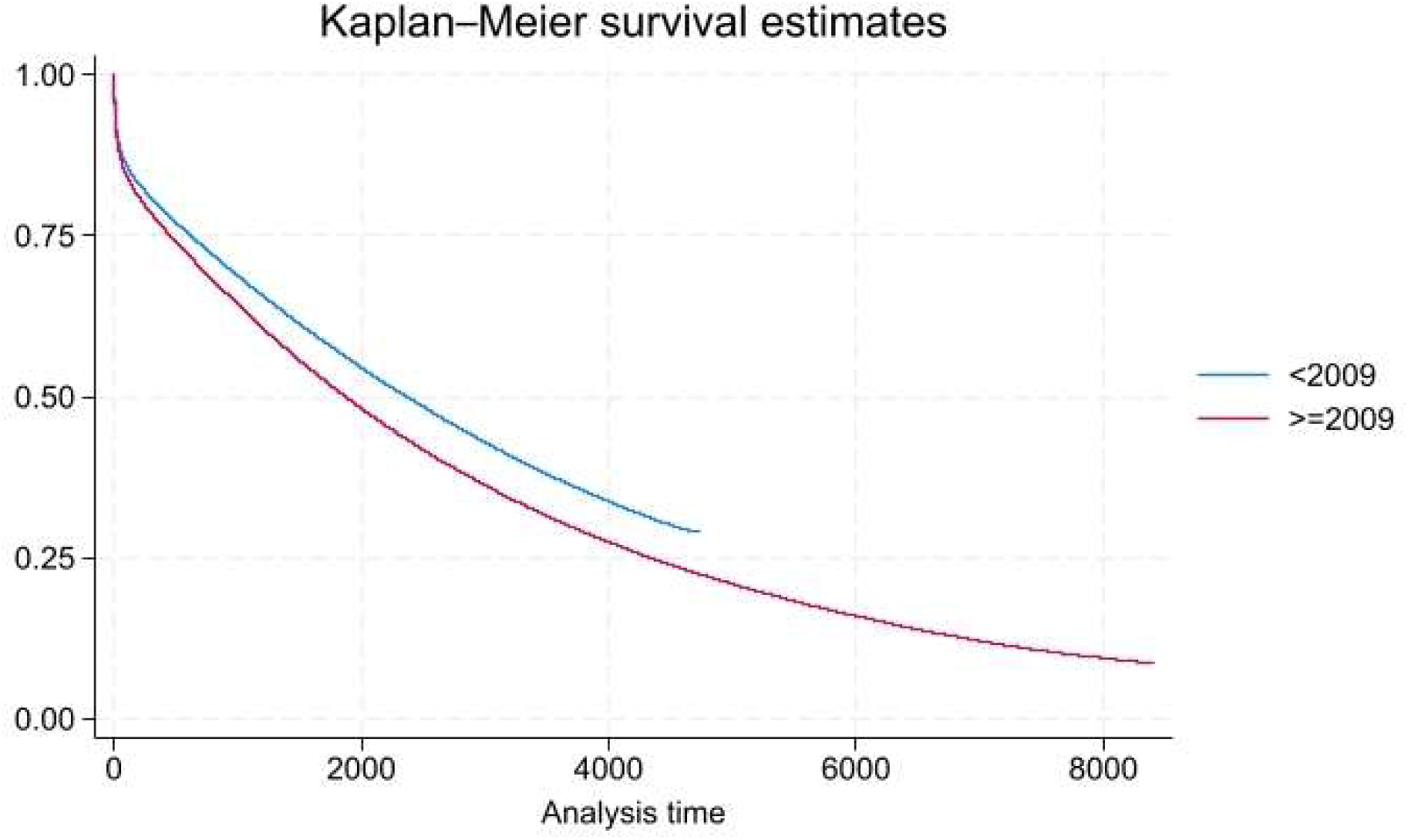
Survival after ischaemic stroke. Survival functions for patients with a first stroke <= 2009 and > 2009. Analysis time in days.

**Figure 2.**
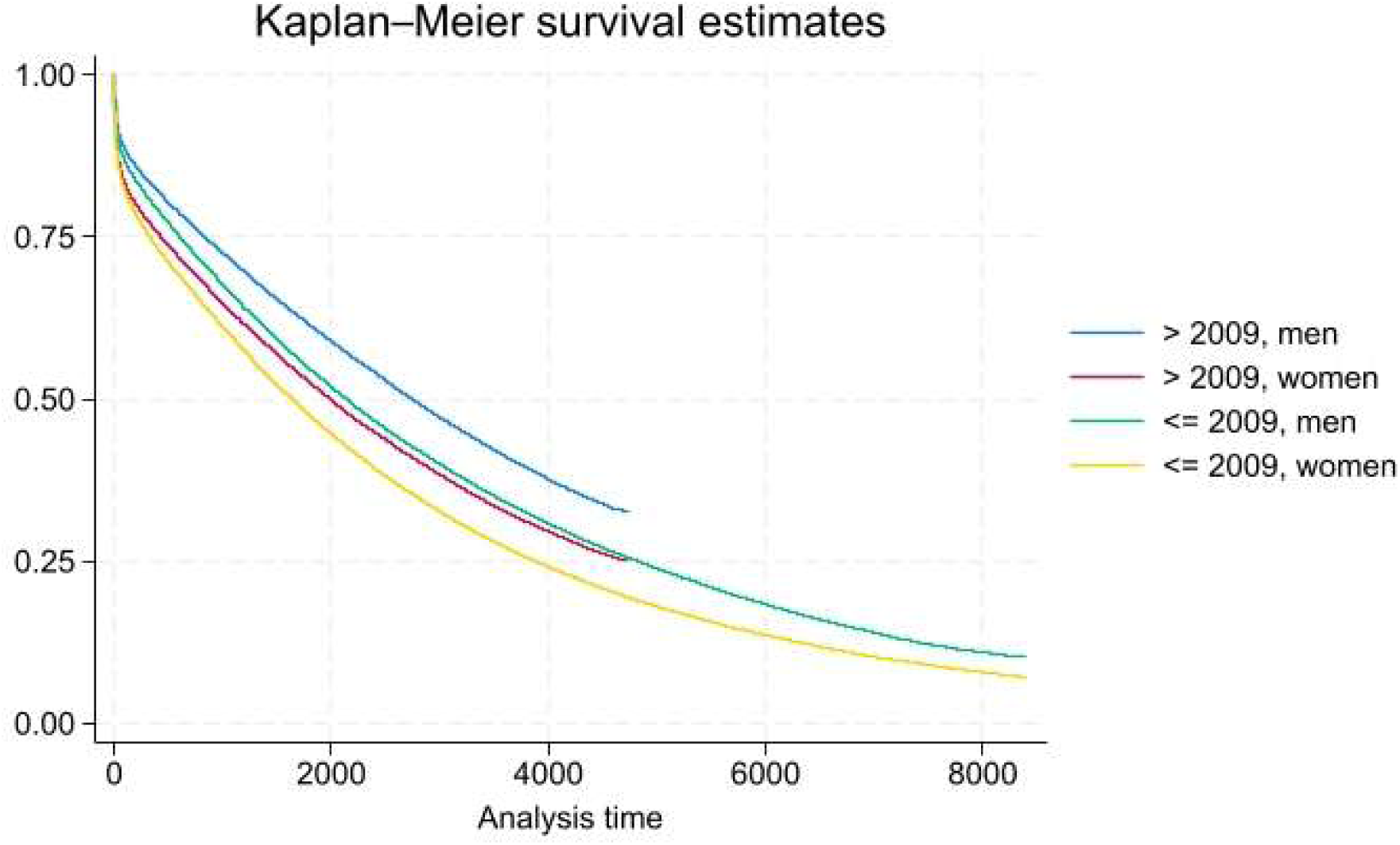
Survival after ischaemic stroke. Survival functions for patient with a first stroke <= 2009 and > 2009, by sex. Analysis time in days.

Figures 3 and 4 illustrates the estimated Kaplan-Meier survival functions for men and women, respectively, in each case stratifying by time-period and educational level. Inspecting the graphs of the survival functions suggests that survival increased between the first and the second time-period for both sexes among patients with at least high school education, but among patients with only primary school only male patients show an improved survival from first to second time-period. Cox regressions yield the following estimates, comparing time-periods separately for the two educational levels: Men: HR low education 0.93, (95% CI: 0.92-0.95); HR high education 0.79, (95% CI: 0.78-0.80); women: HR low education 1.06, (95% CI: 1.04-1.07); HR high education 0.78, (95% CI: 0.76-0.79). Thus, for both sexes, estimated survival for patients with education beyond high school is higher in the second period than in the first period. In contrast, only male patient with high school education (or less) were found to have a higher survival in the second period.

**Figure 3.**
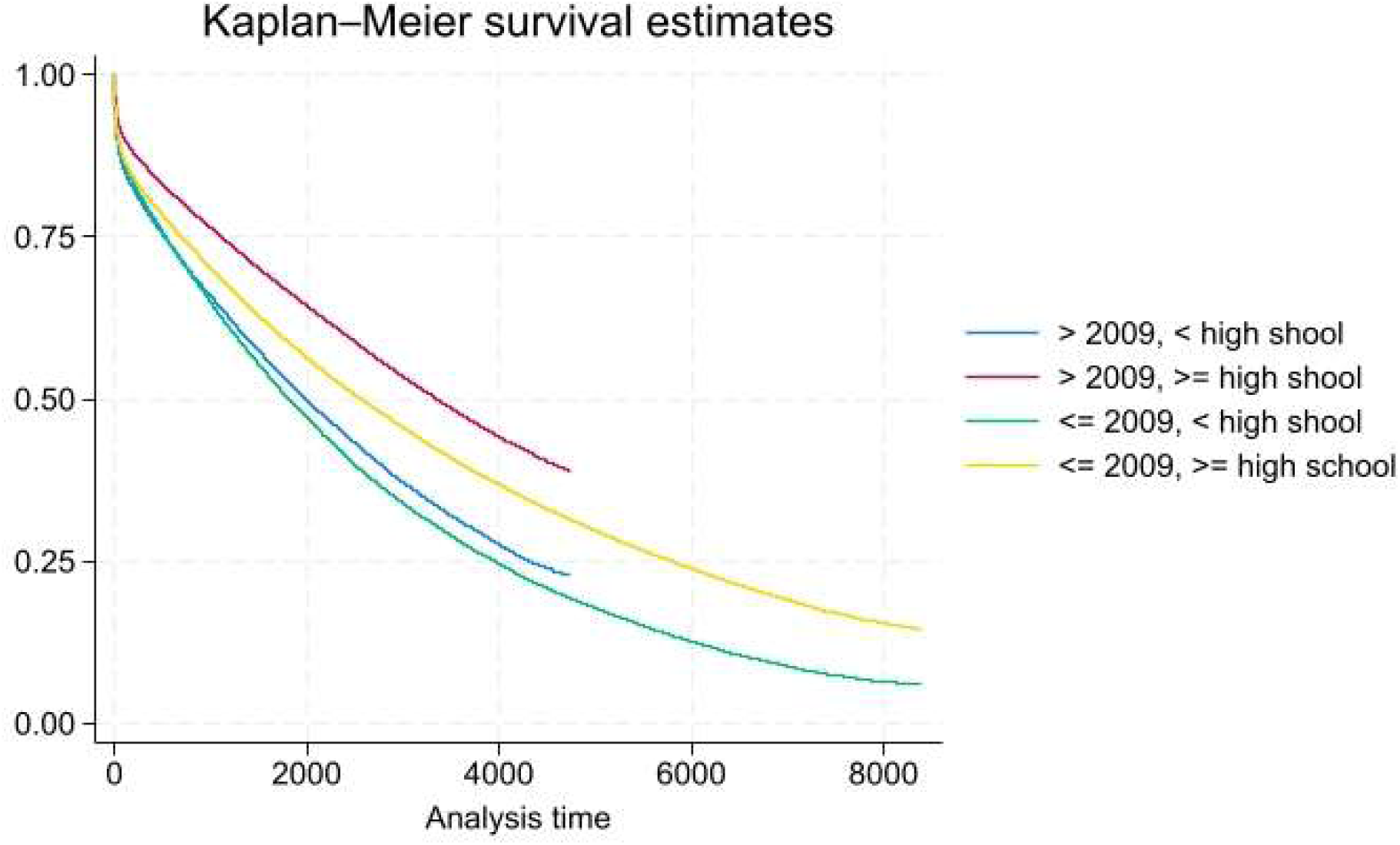
Survival after ischaemic stroke. Patient with a first stroke <= 2009 and > 2009, by sex and educational level. Analysis time in days. Men.

**Figure 4.**
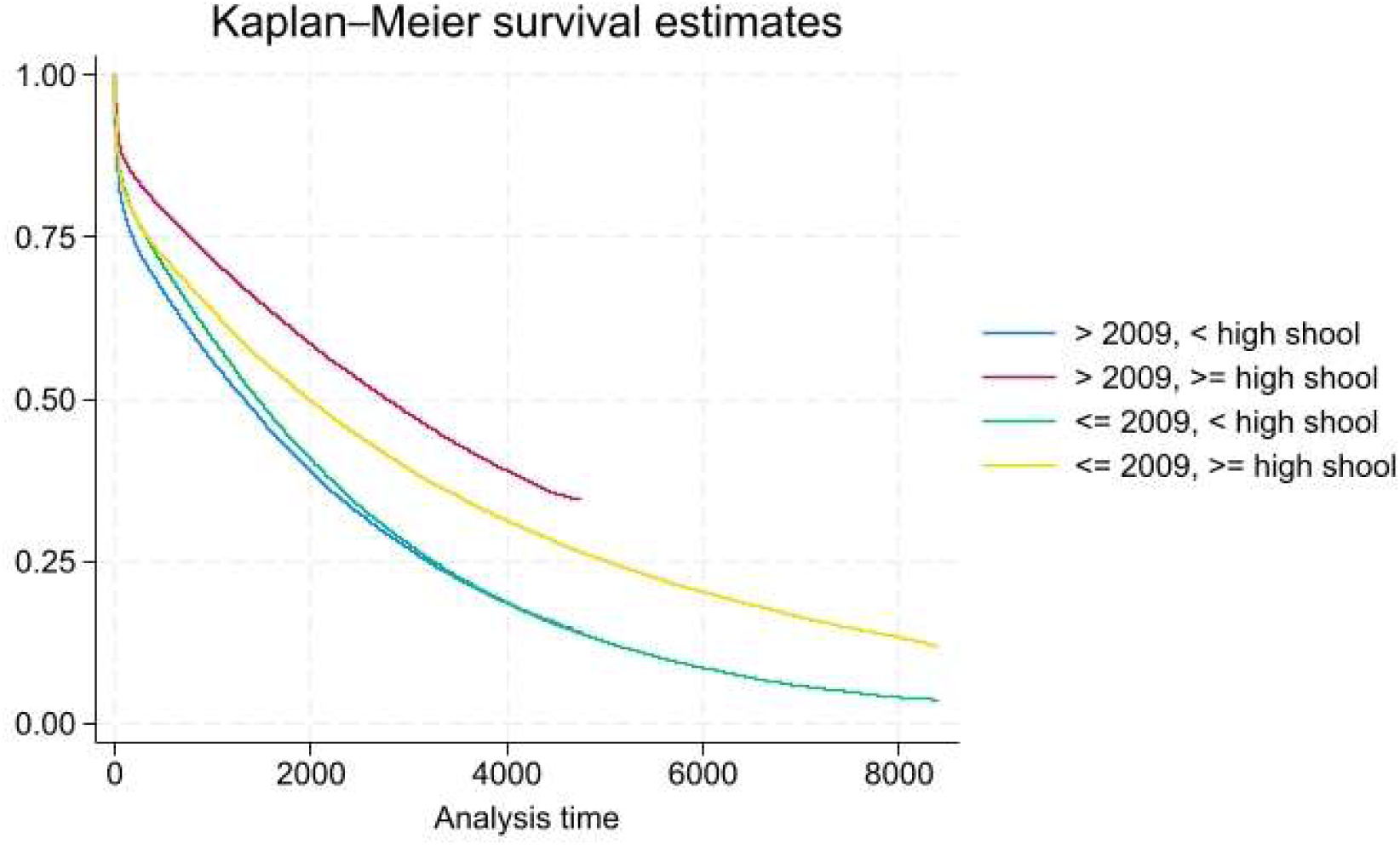
Survival after ischaemic stroke. Patient with a first stroke <= 2009 and > 2009, by sex and educational level. Analysis time in days. Women.

### Change in survival over time – hemorrhagic stroke

Figures 5 – 8 illustrate the survival functions corresponding to Figures 1 – 4 but for patients with a first-time (observed) hemorrhagic stroke. Figure 5 illustrates estimated Kaplan-Meier functions comparing patients with a first hemorrhagic stroke in the first time-period with patients in the second period. The graphs of the functions seemingly largely overlap (log-rank test *p* = 0.51). Thus, on an aggregate level there is no difference in survival between first- and second period patients. Figure 6 illustrates the survival functions when the sample is stratified by sex. The survival functions for men and women, respectively, comparing survival between the two periods do not reveal any differences between survival (log-rank tests: *p* = 0.65; *p* = 0.10).

**Figure 5.**
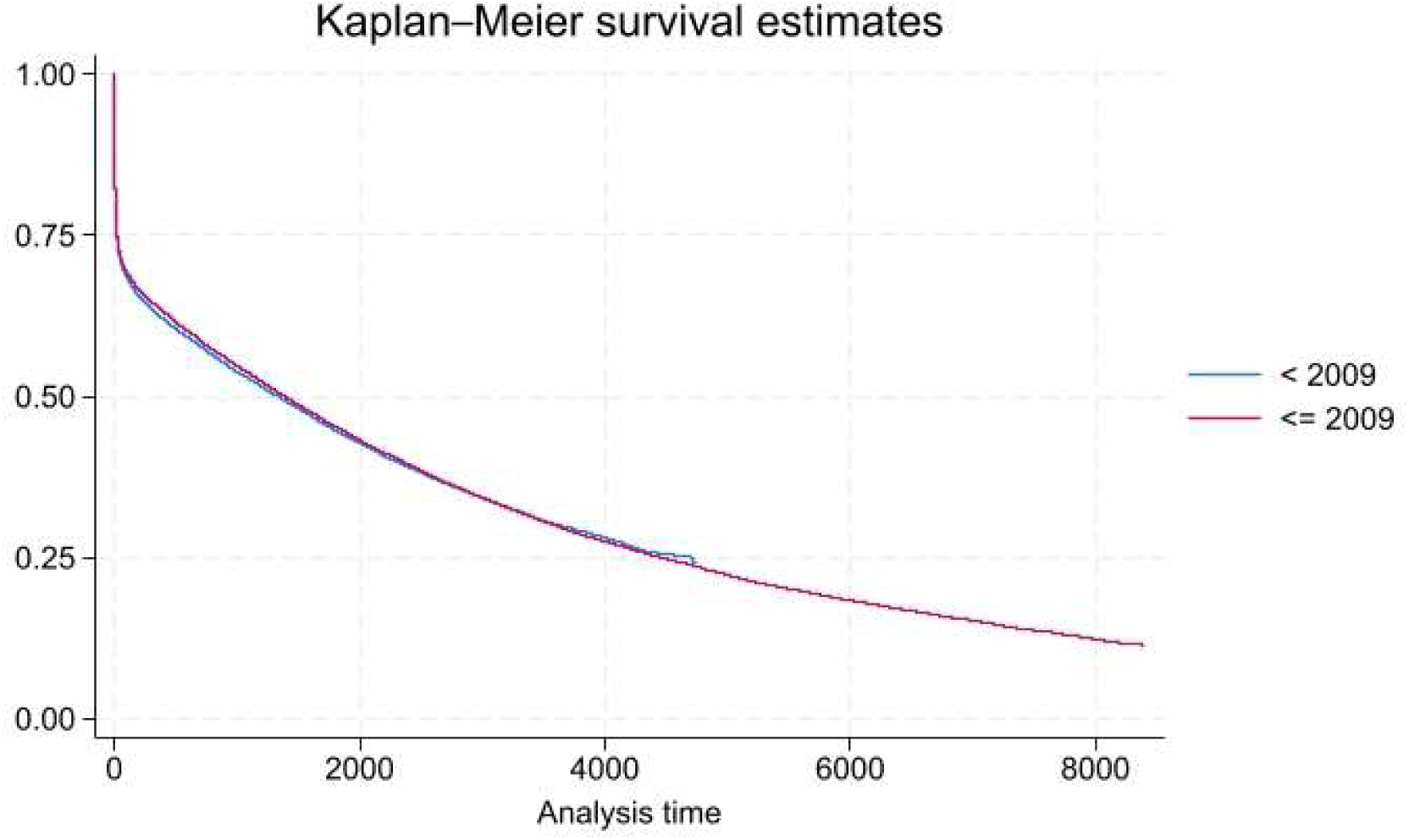
Survival after hemorrhagic stroke. Survival functions for patients with a first stroke <= 2009 and > 2009. Analysis time in days.

**Figure 6.**
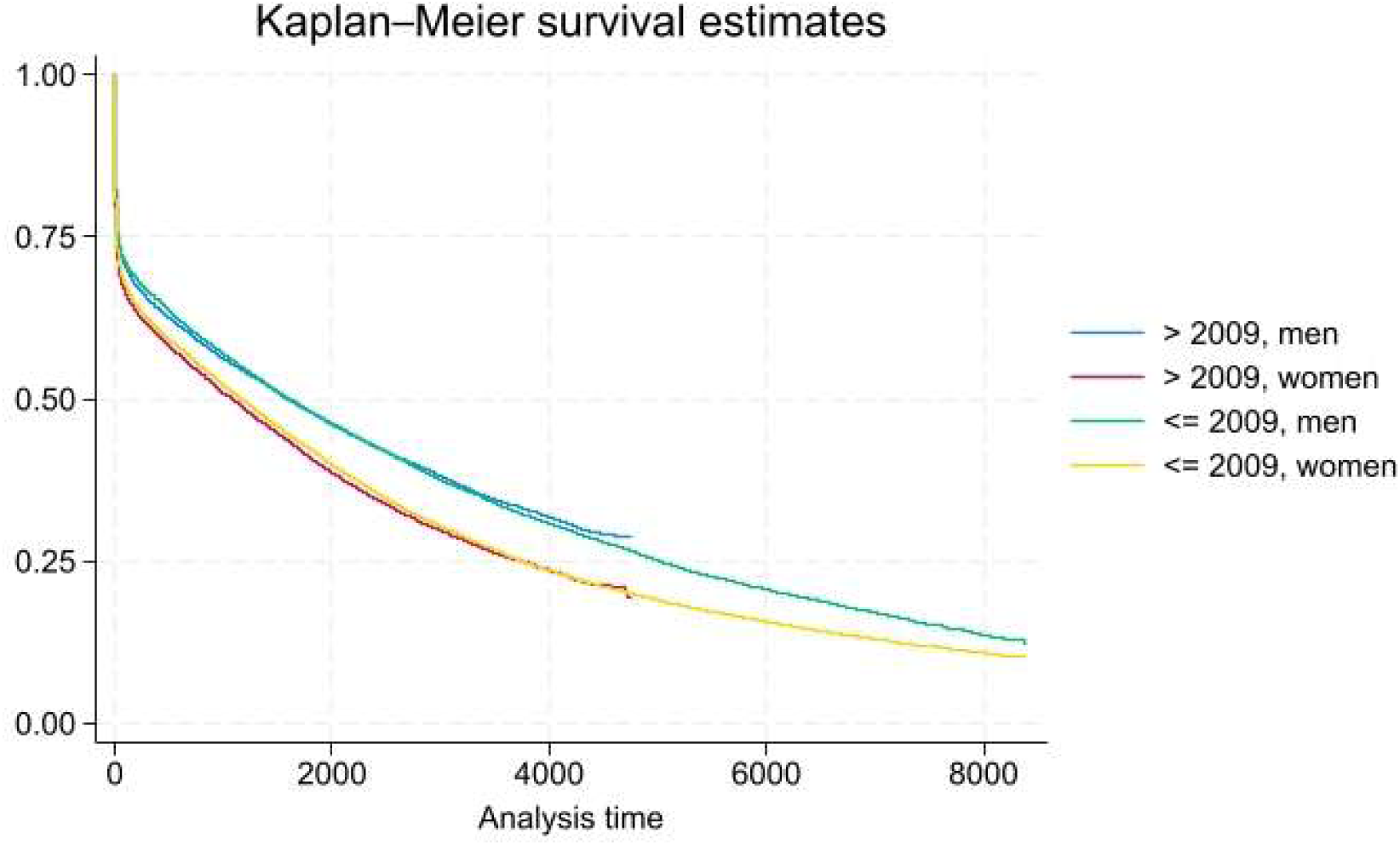
Survival after hemorrhagic stroke. Survival functions for patient with a first stroke <= 2009 and > 2009, by sex. Analysis time in days.

Figures 7 and 8 report the graphs of the survival functions stratifying by time-period and educational level. For both men and women, survival among patients with education beyond primary school did not improve from the first to the second time-period (women, log-rank test *p* = 0.30) or improved slightly (men, log-rank test *p* = 0.01); HR men: 0.96 (95% CI: 0.92-0.99). The corresponding estimates pertaining to patients with lower education reveal that, for both men and women, survival decreased from the first to the second time-period: HR men 1.13, 95% CI: 1.09-1.18; HR women 1.18, 95% CI: 1.14-1.23.

**Figure 7.**
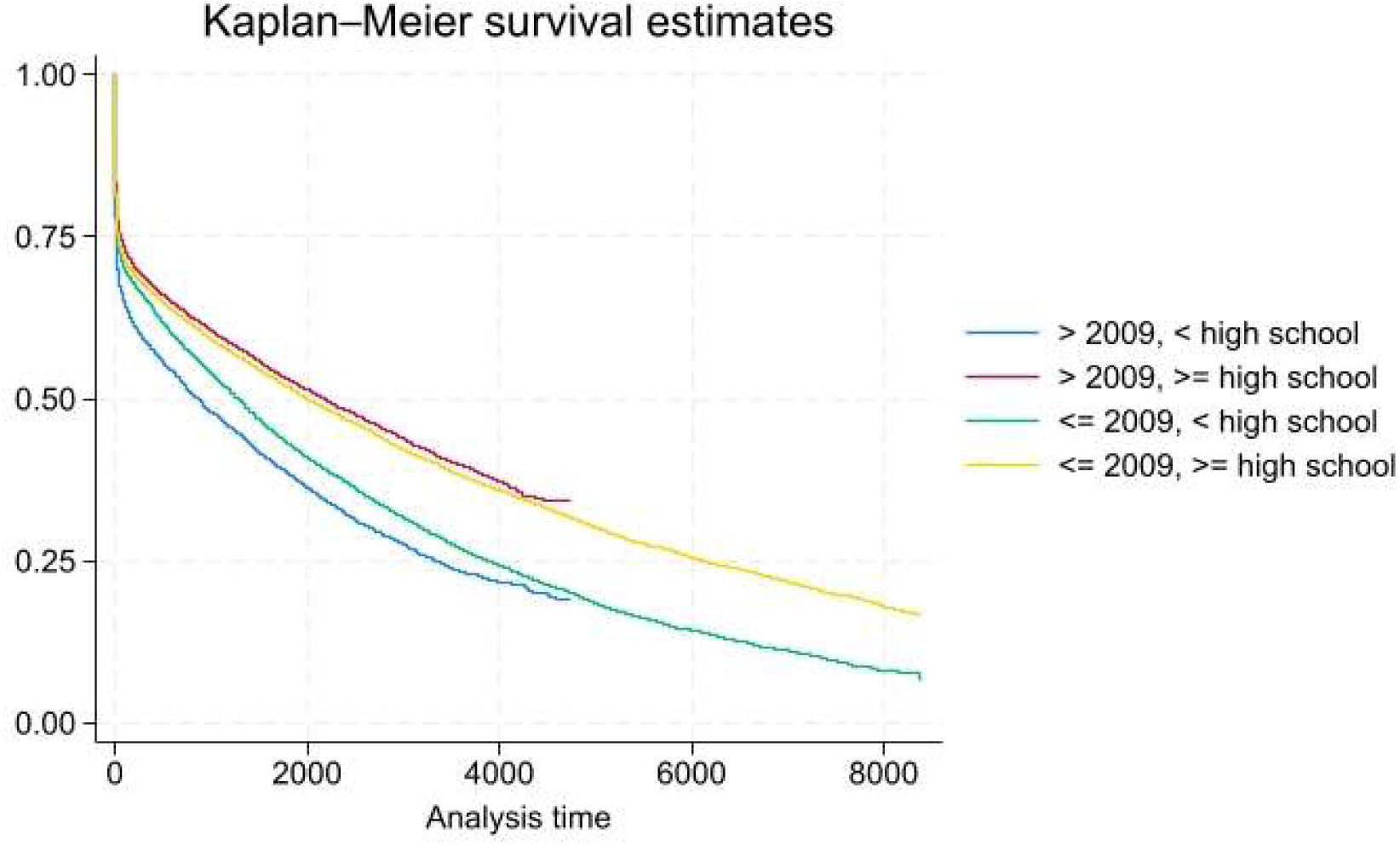
Survival after hemorrhagic stroke. Patient with a first stroke <= 2009 and > 2009, by sex sex and educational level. Analysis time in days. Men.

**Figure 8.**
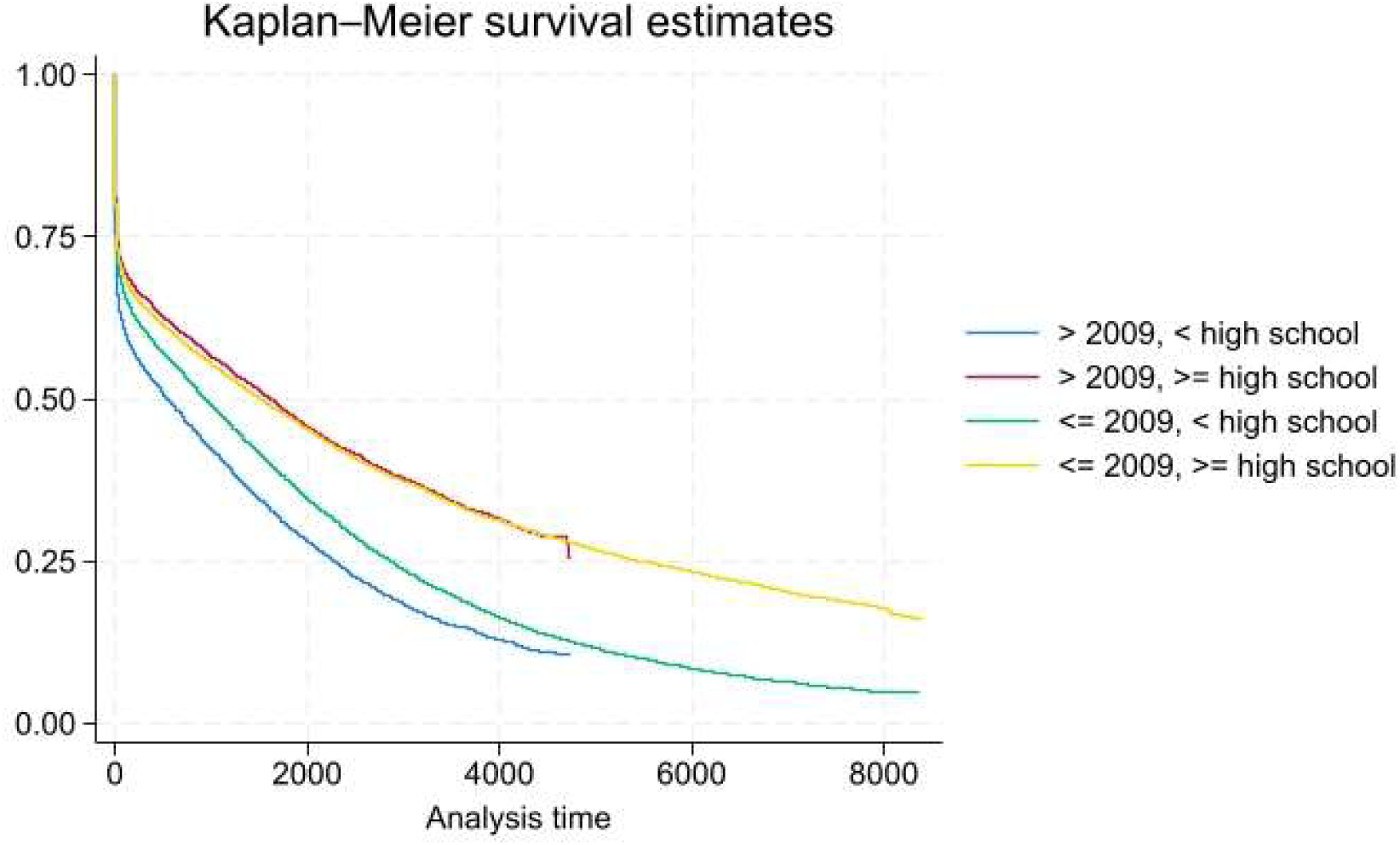
Survival after hemorrhagic stroke. Patient with a first stroke <= 2009 and > 2009, by sex and educational level. Analysis time in days. Women.

### Further stratification, by age

Table 2 reports estimated hazard ratios for stroke occurring 2000-2009 vs stroke occurring 2010-2022 separately for men, women, educational level and age. *First*, note that when stratifying by age the previous conclusion that ischaemic stroke survival among women with low education decreased only remains for patients aged >= 80 years of age, while survival among women aged 50 – 80 increased, and survival among women < 50 years of age did not change. Ischaemic stroke survival among men aged < 50 did not change between time-periods. *Second*, hazard ratios for ischaemic stroke patients with high school education or beyond remained qualitatively the same when stratifying by age: hazard ratios for every age group were estimated < 1. Comparing confidence intervals to infer differences between hazard ratios shows that survival among men aged < 50 improved more than survival among men >= 50, while women aged < 50 improved more only compared to women aged >=80.

**Table 2.** Hazard ratios 2000-2009 vs 2010-2022. By type of stroke, age, and educational level.

|  |  |  |  |
| --- | --- | --- | --- |
| Age group |  |  |  |
|  | < 50 | <= 50 - < 80 | >= 80 |
| Ischaemic stroke, men |  |  |  |
| *Less than high school* | 0.91; 95% CI: 0.69-1.21 | 0.85; 95% CI: 0.83-0.87 | 0.95; 95% CI: 0.93-0.97 |
| *High school or beyond* | 0.63; 95% CI: 0.53-0.74 | 0.80; 95% CI: 0.78-0.82 | 0.83; 95% CI: 0.81-0.85 |
| Ischaemic stroke, women |  |  |  |
| *Less than high school* | 0.81; 95% CI: 0.55-1.20 | 0.88; 95% CI: 0.86-0.91 | 1.06; 95% CI: 1.05-1.08 |
| *High school or beyond* | 0.71; 95% CI: 0.58-0.88 | 0.82; 95% CI: 0.80-0.85 | 0.80; 95% CI: 0.78-0.82 |
| Hemorrhagic stroke, men |  |  |  |
| *Less than high school* | 0.90; 95% CI: 0.66-1.22 | 1.06; 95% CI: 1.00-1.12 | 1.12; 95% CI: 1.05-1.19 |
| *High school or beyond* | 0.76; 95% CI: 0.64-0.91 | 0.93; 95% CI: 0.89-0.97 | 0.98; 95% CI: 0.92-1.04 |
| Hemorrhagic stroke, women |  |  |  |
| *Less than high school* | 0.95; 95% CI: 0.60-1.50 | 1.06; 95% CI: 1.00-1.14 | 1.17; 95% CI: 1.11-1.22 |
| *High school or beyond* | 0.97; 95% CI: 0.77-1.23 | 1.0; 95% CI: 0.945-1.06 | 0.94; 95% CI: 0.89-0.99 |

*Third*, stratifying by age shows that the previous finding that hemorrhagic stroke survival has not increased from the first to the second time-period among patients with high school education or beyond remains only for men aged >= 80 and for women aged < 80. For the other age groups survival did increase from the first to the second time-period. *Fourth*, hemorrhagic stroke survival among patients with education below high school decreased among both men and women aged >= 50, while the survival among younger patients did not change (HR within 95% CI).

## DISCUSSION AND CONCLUSION

In this study the trend of long-term survival after ischaemic and hemorrhagic stroke was estimated comparing stroke occurring in two subsequent time-periods, 2000-2009 vs 2010-2022, using total-population Swedish register data of hospital admissions (main diagnosis and date of admission), mortality and attained education level (at the date of stroke). Comparing survival between the time-periods without controlling for any patient characteristics, show that, whereas survival after ischaemic stroke improved from the first to the second period, no similar improvement could be detected for hemorrhagic stroke patients. This picture remains the same when survival is analyzed separately for men and women (or when sex is controlled for in Cox regressions). It is noteworthy that for ischaemic stroke patients, the hazard ratio for men is lower (statistically significant) than the corresponding ratio for women, and that there is no corresponding difference between hazard rates for hemorrhagic stroke patients. Thus, the survival after ischaemic stroke improved more for men than for women between the first and the second time-period. The explanations for this can be either that there are differences so between men and women when it comes to access to new and improved treatments, or that changes in the characteristics of the male and female populations account for observed changes in survival (or both).

Education is part of what is frequently referred to as human capital (Becker, 1964). There is ample evidence that there exist considerable gradients between, on the one hand, health state and health-related behavior, and, on the other hand, attained education (see e.g., Conti et al., 2010; Clark and Royer, 2013). So, differences in educational level will reflect disparities in both general health state (prior to stroke) as well as lifestyle factors, both plausibly influencing stroke survival. Our findings that the mortality rate following a stroke is lower among patients with highs school education than among patients with less than high school education capture disparities in pre-existing health states and possibly differences in lifestyle factors post stroke. Moreover, we have not investigated if there are any treatment differences between educational groups (reperfusion therapy, oral anticoagulants). Such differences would potentially contribute to explaining the results and should be considered in future research.

## Standard Protocol Approvals, Registrations, and Patient Consents

This study has been approved by the Swedish Ethical Review Authority (reference number: 8866/2023; date of approval: 2024-02-01).

## Data Availability

Data was based on Swedish national registers and individual level data cannot be shared due to national regulations. Original data are available upon application to the relevant authorities.

## Author contributions

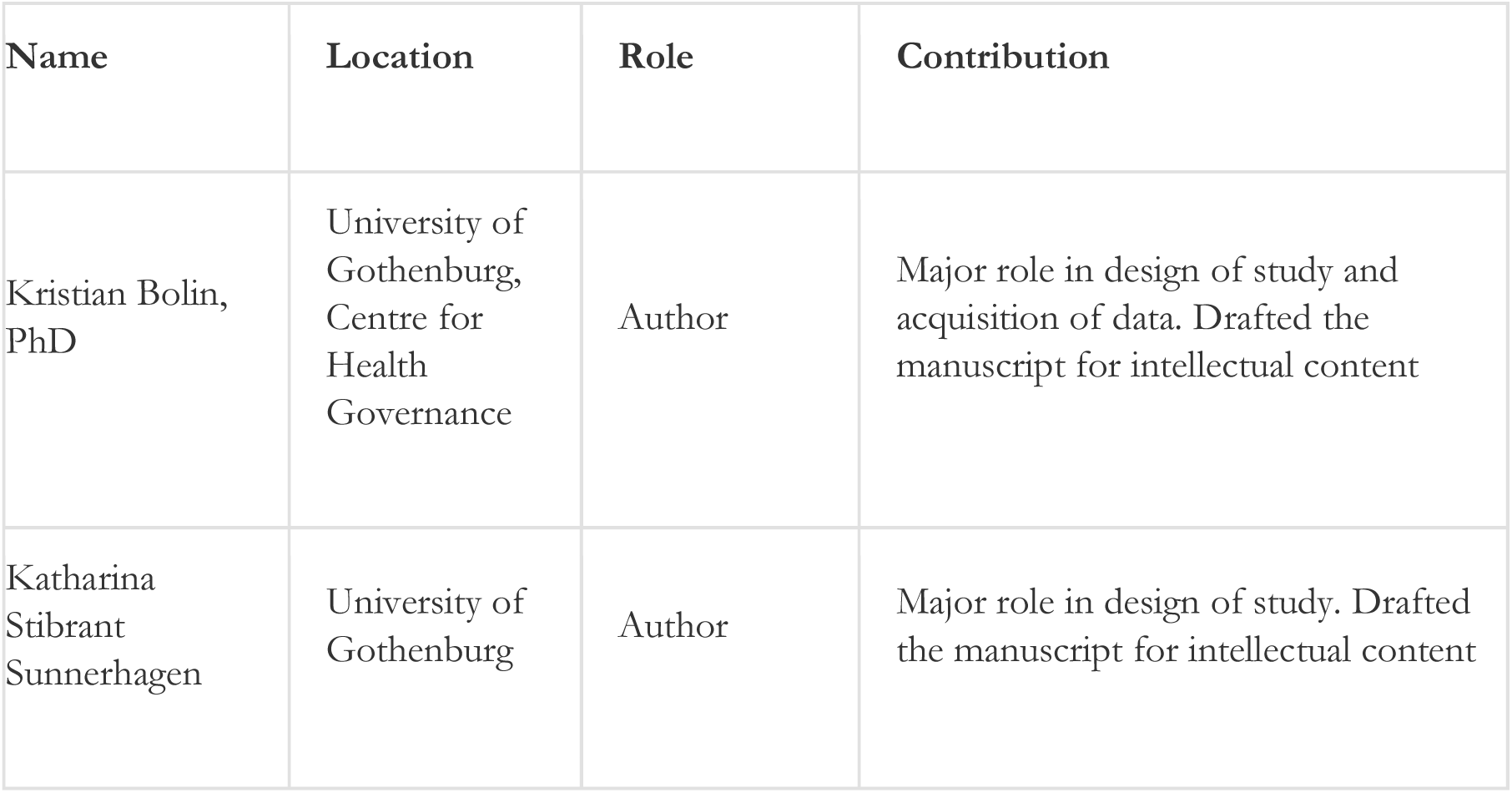

